# Using Mobile Phones For Computer Aided Detection Of Anomalies On Chest Radiographs: Proof Of Concept

**DOI:** 10.1101/19012146

**Authors:** Steven Mitini-Nkhoma

## Abstract

Mastering the art of interpreting chest radiographs can be a long and daunting process. We trained a neural network to classify photographs of chest radiographs into 6 groups. The trained model is hosted on a webserver. The model receives and processes photographs of chest radiographs taken using mobile phones. The system then informs the user the most probable diagnosis. Such systems could prove useful in low resource settings which do not have enough highly trained medical personnel and cannot afford the new generation of CAD based radiology equipment.

## I. INTRODUCTION

In 1895, William Conrad Röntgen serendipitously discovered a previously unknown form of electromagnetic radiation, which later became known as X-rays [1]. He was quick to notice that X-rays were able to penetrate some objects including human soft tissues. Later that year he produced a radiograph – an image formed on a sensitive surface by exposure to X-rays or other forms of radiation – of his wife’s hand [1], [2]. As was demonstrated on the radiograph, X-rays differentially penetrate different materials. A composite object placed between an X-ray source and a detector casts shadows of varying intensities depending on the radiopacity of its components. The medical profession quickly realized how the newly discovered electromagnetic waves could revolutionize the diagnosis and management of fractures [3]. Soon afterwards, portable X-ray machines were used on battle fields to assist surgeons locating and extracting bullets from soldiers [3]. The use of X-rays for diagnosing lung diseases did not lag far behind, and by the 1930s there were X-ray based mass TB screening campaigns [4].

Interpreting chest radiographs is often a formidable endeavor for medical students, junior doctors and sometimes not so junior doctors. As Alexander observes, there is often both inter-observer and intra-observer variations in interpreting chest radiographs [5]. Over the last 3 decades, there has been a shift towards Computer Aided Diagnosis (CAD) in all areas of human medicine. Automation of interpretation of radiographs often relies on machine learning algorithms to either identify an image as belonging to a particular class, or to localize and identify anomalies on radiographs [6].

X-ray machines with integrated CAD systems are being slowly rolled out into health systems all over the world, but the transition has been particularly slow in developing countries, at least in part due to the high cost of the new machines. To address this problem, we built a mobile phone based platform for machine learning aided classification of X-ray images. The platform can be seamlessly integrated into clinical practice without the need for purchasing any specialized equipment.

## II. TRAINING DATA SET

We used Yash Prakash’s Chest X-Rays dataset to train a neural network to interpret chest radiographs [7]. The dataset contains de-identified radiographs grouped into folders depending on the diseases portrayed on the images. The 6 folders are named Atelectasis, Infiltration, Effusion, Cardiomegaly, Fibrosis and Normal. Figure 1 below shows a sample of the images from the dataset.

**Figure 1:**
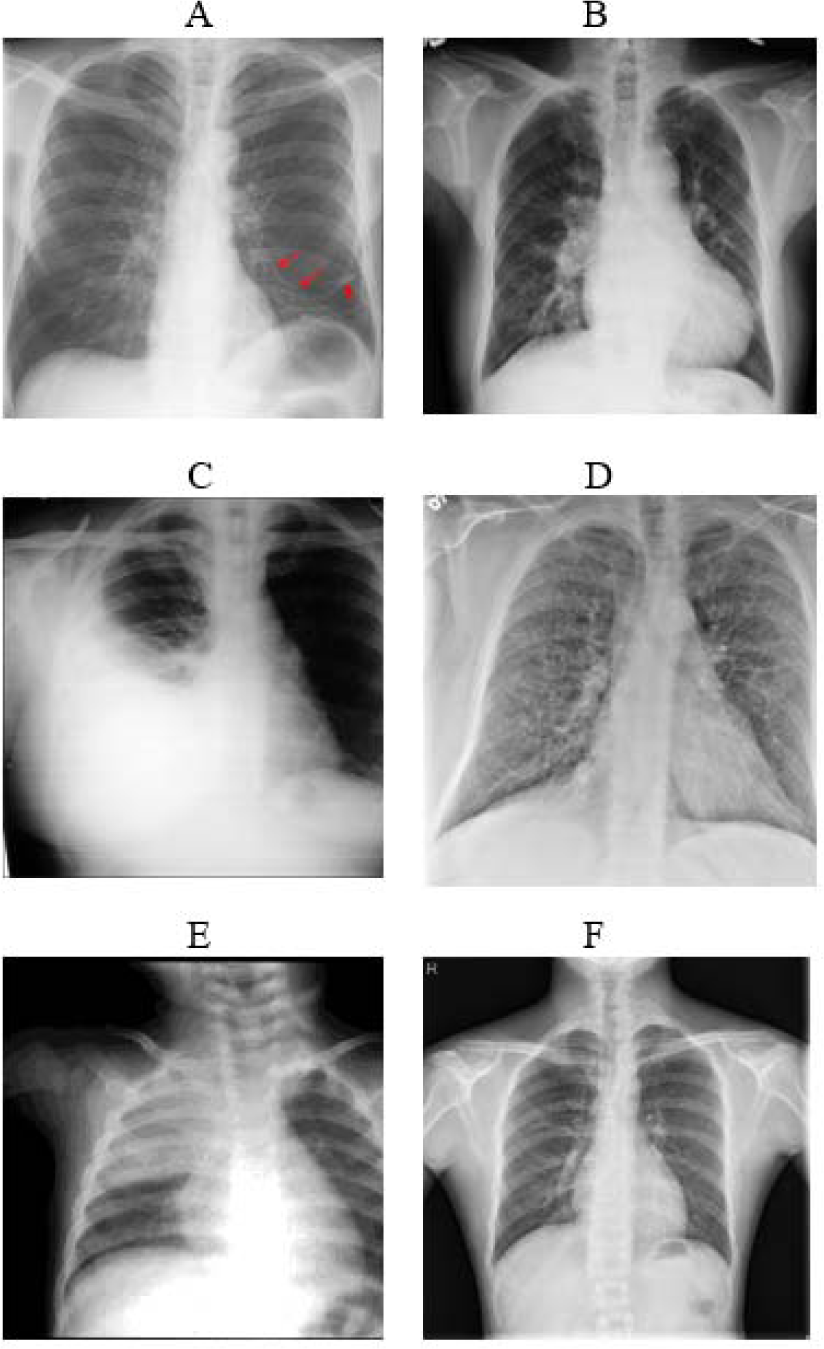
Images from the Chest X-Rays dataset depicting atelectasis (A), cardiomegaly (B), effusion (C), fibrosis (D), consolidation (E) and absence of anomalies(F)

## III. IMAGE PRE-PROCESSING

A medically trained professional re-examined all images in Prakash’s dataset to ensure that they had all been placed into the right class. To expand the dataset, we rotated the images to varying degrees. As the images in Prakash’s dataset come in different sizes and formats, we converted all the images into 600 × 600 pixel JPEG files.

## IV. MACHINE LEARNING PLATFORM

We used Nanonets to build as well as deploy our radiograph classification model [8]. Nanonets is a web based service designed to simplify the training as well as deployment of machine learning models. Creating classification models using Nanonets is a 2 step process; the user uploads the training set into different folders according to their classes, and then invokes the TRAIN command to create a classification model from the data.

For the front end, we created an android application using MIT App Inventor© [9]. The application takes a photograph of a radiograph, converts it into a 600 × 600 pixel JPEG and then sends it to the online machine learning model for analysis.

The model sends back probabilities of the image falling into each of the given categories.

## V. RESULTS AND DISCUSSION

Our model achieved 73% accuracy when tested on a validation data set. The model achieved 70% accuracy when tested under real life situations, using the mobile application running on a Samsung Galaxy A7 2016 to acquire the images. The model was particularly poor at distinguishing images belonging to the Cardiomegaly and Effusion classes as shown in Figure 2 below.

**Figure 2:**
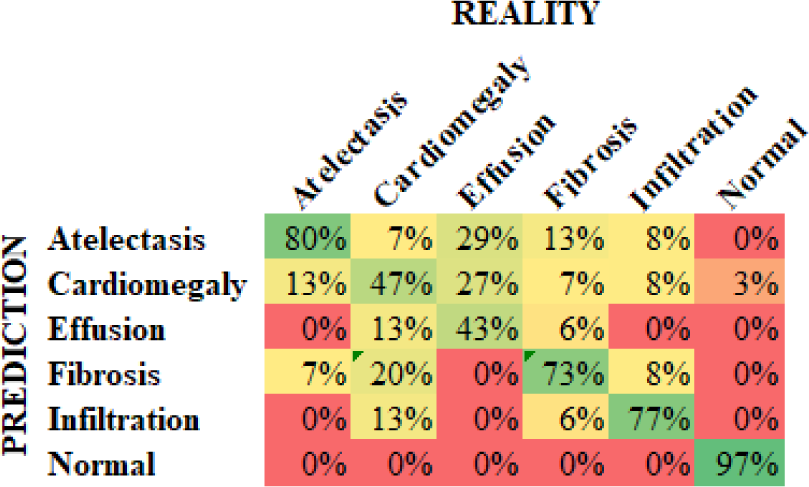
Confusion matrix of the chest radiograph classification model

Cardiomegaly is the enlargement of the heart while a pleural effusion is when there is fluid in the space surrounding the lung [10]. Cardiomegaly often causes an effusion. A large effusion obscures the heart on a radiograph, making it difficult for one to examine the size of the heart shadow as shown in Figure 3 below [11]. The presence of cardiomegaly will instantly alert the doctor to look for an effusion in the image, and vice-versa [11]. The inability of the system to consistently tell the two phenomena apart is therefore unlikely to cause confusion in the real world.

**Figure 3:**
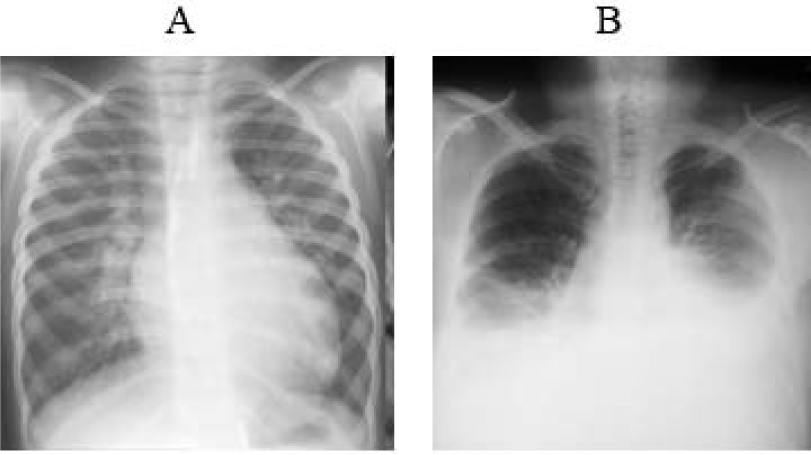
Images from the Chest X-Rays dataset depicting cardiomegaly (A) and an effusion obscuring the heart shadow (B and C)

## VI. CONCLUSION

We have created a mobile platform powered by machine learning than can distinguish radiographs belonging to 6 classes. Platforms like ours can help simplify interpretation of chest radiographs, and would be particularly useful in low resource settings where radiographers are inadequate. Such platforms would also make excellent learning tools for medical students.

## Data Availability

The data refered to in the manuscript is available from the author upon request.

## REFERENCES

[1] R. F. Mould, A History of X-rays and Radium: With a Chapter on Radiation Units, 1895-1937. IPC Building & Contract Journals, 1980.

[2] A. Hessenbruch, “A brief history of x-rays.,” Endeavour, vol. 26, no. 4, pp. 137–41, Dec. 2002.

[3] A. B. Reed, “The history of radiation use in medicine,” Journal of Vascular Surgery, vol. 53, no. 1 SUPPL. Mosby Inc., pp. 3S–5S, 2011.

[4] J. E. Golub, C. I. Mohan, G. W. Comstock, and R. E. Chaisson, “Active case finding of tuberculosis: historical perspective and future prospects [Review Article].”

[5] K. Alexander, “Reducing error in radiographic interpretation.,” Can. Vet. J. = La Rev. Vet. Can., vol. 51, no. 5, pp. 533–6, May 2010.

[6] H. Abe et al., “Computer-aided Diagnosis in Chest Radiography: Results of Large-Scale Observer Tests at the 1996–2001 RSNA Scientific Assemblies,” RadioGraphics, vol. 23, no. 1, pp. 255–265, Jan. 2003.

[7] Y. Prakash, “Chest X-Rays Dataset.” 2017.

[8] Nano Net Technologies Inc, “Nanonets.” 2019.

[9] E. W. Patton, M. Tissenbaum, and F. Harunani, “MIT App Inventor: Objectives, Design, and Development,” Comput. Think. Educ., pp. 31–49, 2019.

[10] V. Nguyen, “Dilated Cardiomyopathy: Practice Essentials, Background, Pathophysiology,” 2018. [Online]. Available: https://emedicine.medscape.com/article/152696-overview. [Accessed:11-Nov-2019].

[11] V. S. Karkhanis and J. M. Joshi, “Pleural effusion: Diagnosis, treatment, and management,” Open Access Emerg. Med., vol. 4, pp. 31–52, May 2012.

